# Hair Cortisol as a Marker of Physiologic Stress in Residency Training

**DOI:** 10.64898/2026.01.16.26344232

**Authors:** Laura Hinz, Kirstie Lithgow, Kiara Kunimoto, Gregory Kline

## Abstract

**Background:** Hair cortisol analysis allows assessment of long-term cortisol exposure and may provide insight into chronic hypothalamic-pituitary-adrenal activation in medical residents and residency on-call responsibilities.

**Objective:** To determine the hair cortisol concentration(HCC) representing 3 months of medical residency and secondarily, its association with various on-call models (in-hospital, night float, home call and no call).

**Design:** Cross-sectional study of 66 medical residents who were recruited to provide hair samples collected after a three-month block in medical residency.

**Setting:** Academic, tertiary health care system.

**Participants:** Volunteer sample of first through third year medical and primary care residents.

**Exposure:** 3 cm of hair was divided into 3 segments of 1 cm each; each segment represented 1 month of cumulative cortisol production.

**Main Outcome Measure:** HCC results were compared to a published, cortisol assay-specific normative population reference interval. HCC results were interpreted according to a priori categorizations of moderate (+1.5SD), considerable (+2SD) or extreme (> +3SD) HCC elevations. Associations with various on-call models were an exploratory secondary outcome.

**Results:** The median age was 28 (26–30) years with median sleep duration of 2 hours on in-hospital call. 40% of trainees had at least one HCC segment above the threshold deemed marked elevation. Median HCC was significantly higher for in-hospital and night float *vs.* no call (285 ng/g and 335 ng/g vs 78 ng/g p<0.05) and approached significance compared to home call (190 ng/g, p= 0.06).

**Conclusions and Relevance:** We have described chronic exposure to endogenous cortisol in medical residency. Nearly half of trainees experienced at least one month of severe hypothalamic-pituitary-adrenal axis activation in a 3-month timeframe; many had marked chronic cortisol elevations across the entire 3 month observation frame. HCC was higher in months where in-hospital on-call was required. This may have implications for long-term health of trainees and raises questions about the structure of duty hours and sequence of care acuity blocks within residency training programs.

## Introduction

There has long been recognition that the demands of medical training may adversely impact the hypothalamic-pituitary-adrenal (HPA) axis and cortisol production(1). Chronic hypercortisolism and loss of the normal diurnal pattern of endogenous cortisol secretion have been closely associated with multiple metabolic risks including hypertension, insulin resistance, diabetes, bone loss, and psychiatric disturbances (2,3,4). This may occur in the context of neoplastic hypercortisolism as well as non-neoplastic hypercortisolism (formerly called “pseudo-Cushing’s disease”) (5). The adverse health associations of psychological and physiological stress are well described (6) although the biological mediators of stress effect are less certain. Prior studies have suggested a possible role for stress-induced hypercortisolism in the process (7). If confirmed, cortisol measures may serve as either a biomarker of stress-induced health risk with possible prognostic or even therapeutic implications (8,9,10).

Hair cortisol analysis is a novel tool that allows for retrospective, summative analysis of continuous, longer-term endogenous cortisol exposure, with one centimeter of hair representing approximately one month’s exposure duration (11). Unlike other single-point biochemical methods of cortisol measurement (measurement in blood, saliva or urine) which are susceptible to variation due to factors such as acute stress, diurnal rhythm, and pulsatility, hair cortisol provides information about long term endogenous cortisol exposure over months to years (11), similar to the way that hemoglobin A1c estimates the average blood glucose level from over time. One promising application of hair cortisol analysis is for studying exposure to chronic stressors (12,13,14). Elevated hair cortisol levels have been previously demonstrated in young adult shift workers compared with controls (15), and have been associated with burnout symptomatology (16). In addition to the obvious advantage of longer time-related cortisol exposure estimates versus a single point serum cortisol measure, HCC has been shown to be unchanged by other factors such as smoking, body mass index or common drugs including oral estrogens (17).

In response to calls for increased regulation and restriction of resident duty hours as a potentially modifiable major stressor, many medical residency programs have shifted from the traditional 24 hour on-call model towards alternative strategies including shorter shifts, night float, and protected sleep time (18). Numerous studies have assessed the impact of these changes on resident wellbeing, education, sleep, and alertness (16,18–23). However, the physiological impact of these various call models has not been evaluated and no biomarkers yet identified to measure individual resident response to on-call duties that have different anticipated stress effects. Our institution employs a variety of call models, including 26-hour in-hospital call, home call, and night float (scheduled 8-12-hour in-hospital shifts). Accordingly, trainees are exposed to varying call models during different clinical rotations, which are structured in 4-week blocks. We sought to use HCC measures to characterize the degree of chronic, endogenous HPA axis activation in medical residents over a 3-month frame, and the proportion in whom frank hypercortisolism is observed. Secondarily, we evaluated the association of different call models (night float, in-hospital call, home call, and no call) with hair cortisol analysis as one potential major component contributing to the stress of a medical residency.

## Methods

The procedures followed in this study were in accordance with and approved by the Conjoint Health Research Ethics Board at the University of Calgary, Alberta, Canada REB16-1855. We conducted a cross-sectional study of medical trainees enrolled in the internal medicine, and family medicine programs at the University of Calgary over a three-month (3 rotation) period from Jan 2017-May 2018. The trainees in these programs were required to participate in a mixture of four different call types (night float, in-hospital call, home call or no call) depending on the clinical rotation. Additional eligibility criteria included the presence of hair on the top of the head of at least 3 cm in length and willingness to provide a shaved hair sample. Individuals with known diagnosis of cortisol excess or deficiency, or any current use of exogenous corticosteroids were excluded from participation. Each participant provided written consent to participation and collection of the hair sample.

The four different call models were structured as follows: in-hospital call required the trainee to be present in the hospital (“in-hospital”) from 0800 until 1000 hours the following morning; “night float” required the trainee to be in-hospital from 2000 to 0800 hours the following morning (usually 3 or more consecutive shifts with opportunity to rest during daytime hours); “home call” allowed the trainee to leave the hospital after completion of clinical duties with return as needed; “no call” meant the trainee did not have any after-hours clinical duties during the rotation.

Demographic data including age, sex and year of training, was collected from participants at an enrollment visit, prior to the start of a 3-month (3 rotation) training period. In the third week of each four-week rotation, participants were emailed a link for an online survey enquiring about recalled details of their current rotation including clinical rotation, call model, (in-hospital call, no call, night float, home call), number of call shifts scheduled, estimated average hours of sleep per night on call, estimated average hours of sleep per night not on call, and estimated average number of pages per night of call. Participants also completed the Perceived Stress Scale (24) and the Holmes-Rahe Life Stress Inventory (25).

At the second visit, at the end of the three-month study period, we collected hair samples from participants for retrospective hair cortisol analysis. A sample of approximately 100 hairs was trimmed at the vertex of the scalp of each subject using a previously described technique (11). Given one centimeter of hair has been approximated to a one month time frame (11), each three-centimeter segment of hair was correlated with three separate rotations and their respective call models as per participants’ responses to the questionnaire. The scalp terminus of the hair sample was labelled, and all samples were accompanied by information about date of rotations and date of hair collection, then shipped to the reference lab (Clinical Pharmacology Lab, London, ON) for analysis. As previously described by Greff *et al*, the hair was first weighed then washed, then minced into small pieces (26). The cortisol was extracted with organic solvents and evaporated under nitrogen and heat. The sample was resuspended in phosphate buffered saline and analyzed using a commercially available salivary cortisol EIA kit (ALPCO Diagnostics, Salem, NH). Intra-assay variability was previously reported at 5.05% with upper and lower limits of detection of 25 ng/g and 2000 ng/g (27).

### Hypercortisolism in Medical Residents

We utilized the reference population mean hair cortisol concentration (HCC) previously published by Igboanugo *et al*(28) which closely matched a prior study in healthy Spanish volunteers(17); both using the same ALPCO cortisol ELISA for HCC. From that study, the reference population mean(SD) was 128.3 (113.6) ng/g, with “moderate” elevation deemed to be the mean+1.5 SD which is > 298.6 ng/g, “considerable” elevation denoted as the mean + 2SD which is > 355.4 ng/g and “extreme” elevation described as the mean + 3SD which is > 468.9 ng/g. We recorded descriptive HCC results for the study population, in addition to individual HCC analysis to determine the number of subjects with at least one month of moderate or worse hypercortisolism as defined by the categories above. A more longitudinal analysis included estimation of the prevalence and patterns of sustained hypercortisolism across all 3 of the one-month segments, specifically the percentage who never show HCC elevation, those with at least one elevation and those with all 3 segments showing HCC elevation.

#### Hair cortisol levels according to four types of call duty

To ascertain the association between call type and hypothalamic-pituitary-adrenal (HPA) axis activation, hair cortisol results were pooled according to corresponding call type (in-hospital, home, night float, and none) and tested for significant difference across all four with subsequent individual paired comparisons.

### Statistical Analysis

Baseline participant characteristics and survey results were reported as means (SD) or median (IQR) as appropriate to their normal or non-parametric distribution. One-way ANOVA comparison between multiple non-parametrically distributed groups was conducted using the Kruskal-Wallis test with Tukey correction for multiple comparisons. The Mann-Whitney test was used to compare two non-parametrically distributed groups and Spearman correlation for tests of association. For each test, p<0.05 was taken to indicate statistical significance. All statistics were performed with GraphPad Prism 6.0 (GraphPad Software, Boston, USA).

## Results

Overall, 66 participants completed the study, 21 men and 45 women, representing 24% of all residents contacted for potential participation. The median (IQR) age was 28 (26–30) years. Most participants were in their first year of residency (Postgraduate Year 1, i.e. PGY1) (72.7%), with 12.1% in PGY2 and 15.2% PGY3. Three participants (4.5%) reported some form of glucocorticoid use in the past but none during the study period. There were no major life events on the Perceived Stress Scale or Life Stress Inventory that persisted for more than one month. One subject had only 1 usable hair segment, but all others had 3 usable segments, for a total of 196 segments representing 196 months of observation. The most common call type associated with a unique hair segment was in-hospital call (50%), with 30.1% home call, 6.1%-night float, and 13.8% no call.

### Hair cortisol levels

The individual hair segment cortisol data is shown in Figure 1 where it may be seen that there was a very wide range of inter-individual variation. Half of the trainees had at least one hair segment cortisol above the threshold deemed to indicate considerable or extreme elevation (greater than 2-3 SD above the population mean for the one-month frame).

**Figure 1:**
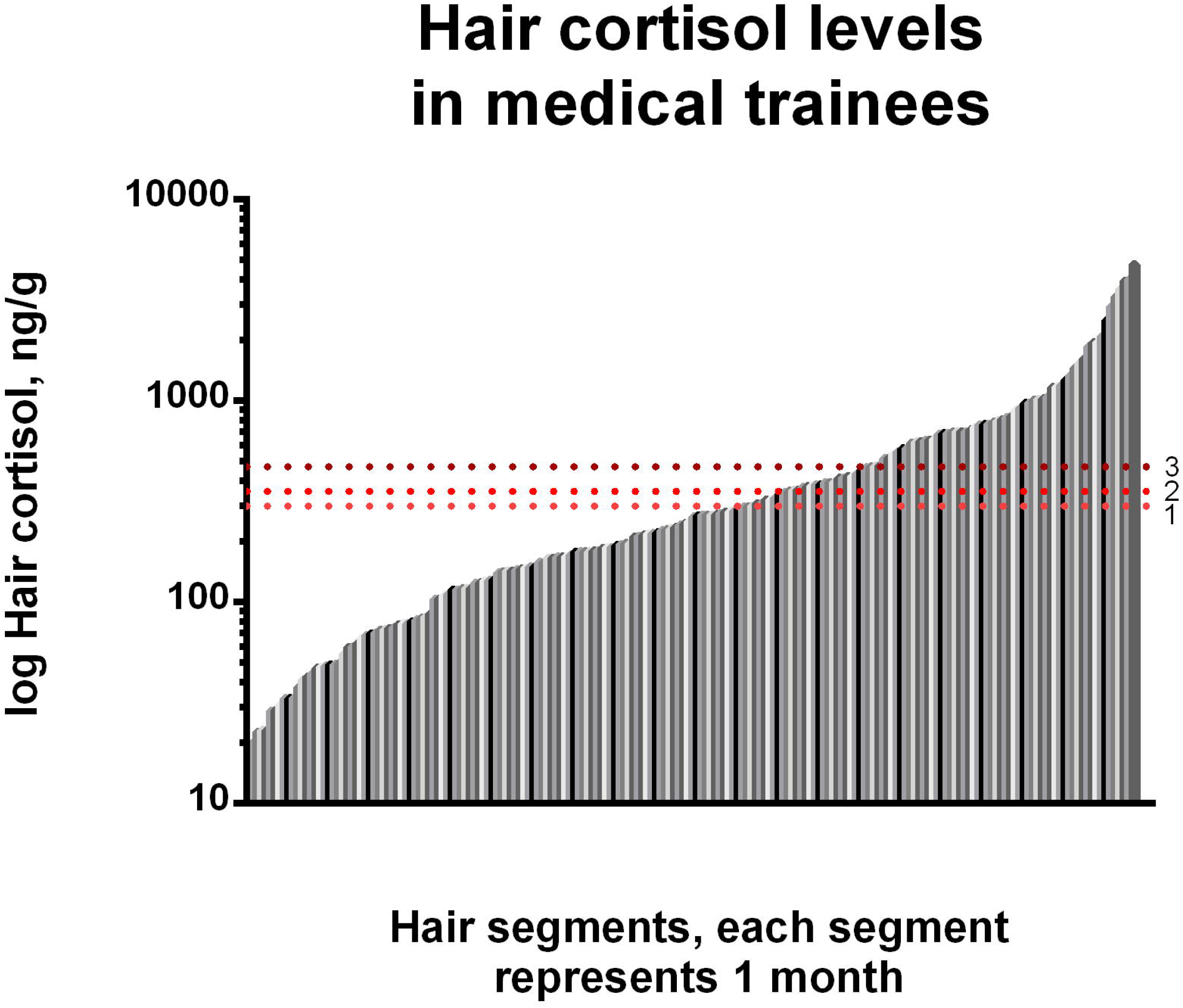
Individual participant level, hair-segment data for unique hair cortisol concentrations, log-scale. Thresholds to determine moderate and marked elevation from Igboanugo (17). 1 = modest elevation (+1.5 SD), 2= considerable elevation (+2 SD), 3 = extreme elevation (+3 SD).

After categorizing participants according to the three patterns of 3-month trends, there were 43.1% of trainees who never showed more than moderate elevation of hair cortisol in any of the 3 month-segments, 40.0% who had at least one extreme elevation (> 469 ng/g) in hair cortisol in 3 months and 16.9% where all 3 segments showed extreme elevations in hair cortisol. The corresponding measures (median, IQR) in each of the groups was 122 (67-186) ng/g, 372 (219-560) ng/g and 1019 (775-1943) ng/g (Figure 2).

**Figure 2:**
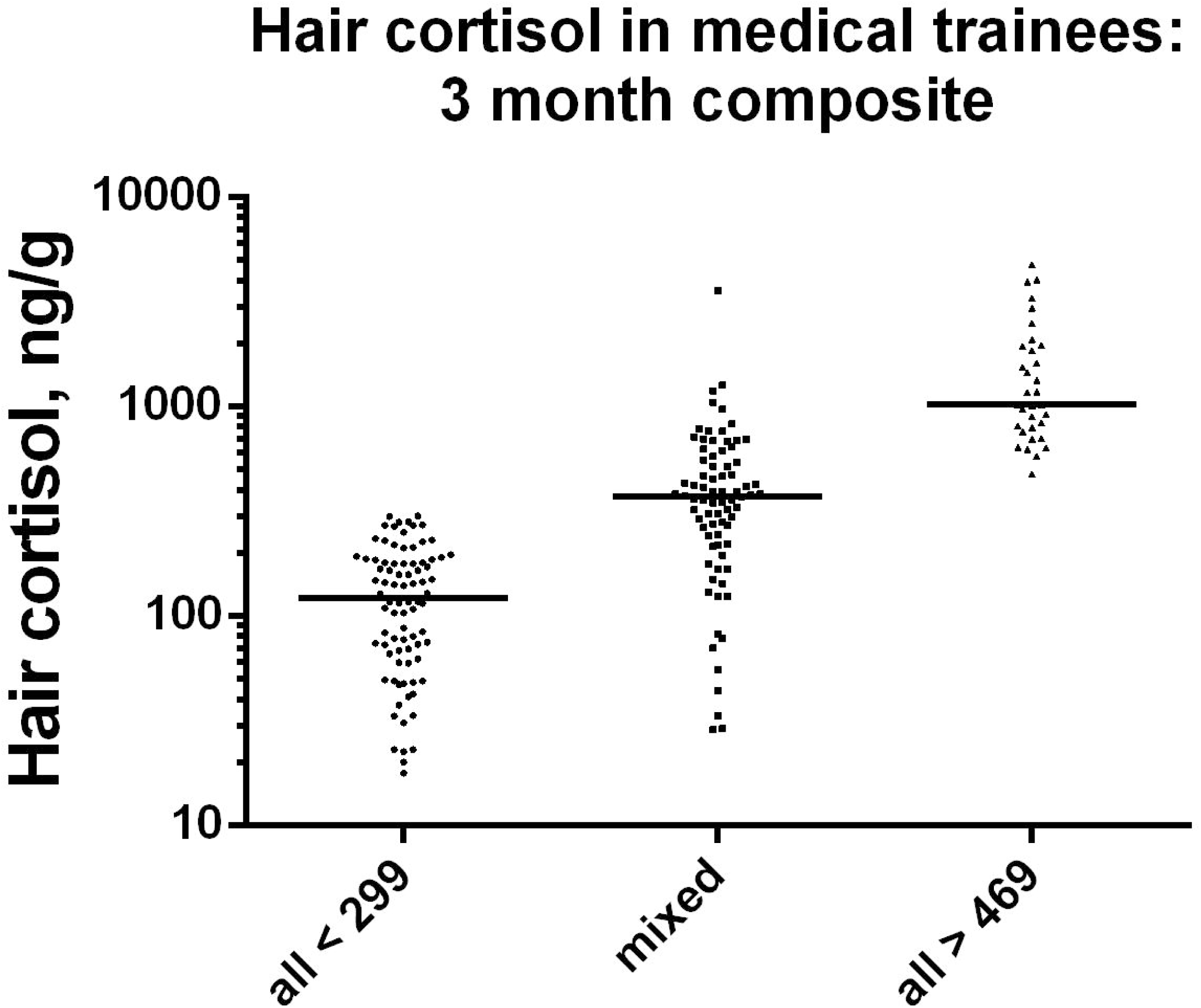
Moderate, Mixed and Sustained, Severe elevations in hair cortisol triads; each point represents one unique subject. Left, subjects who never had a single month-segment > 299 ng/g (modest elevation). Right, subjects with all 3 hair segments showing HCC > 469 ng/g (extreme elevation). Middle, Subjects with at least 1 of 3 month-segments showing HCC > 469 ng/g (mixed elevation).

### Association with on-call type

Table 1 shows the self-reported, recalled within-rotation sleep and interruption rates per call block. Residents reported less sleep on in-hospital call rotations (in-hospital call and night float) with a median of two hours of sleep on in-hospital call and 0.5 hours on night float, *vs*. five hours on home call and six hours on no-call (p<0.0001). Sorted by rotation call-type, the median(IQR) HCC was 285 (128-684)ng/g for in-hospital (n=98 segments), 335 (134-674) ng/g for night float (n=12 segments), 190 (83-422) ng/g for home call (n=59 segments), and 78 (20-220) ng/g for no call (n=27 segments) (Figure 3). There was a significant difference in HCC across the 4 call types (p=0.19, ANOVA). In pairwise comparisons, median HCC was significantly higher for in-hospital *vs.* no call (p<0.05) and approached significance compared to home call (p 0.06).

**Figure 3:**
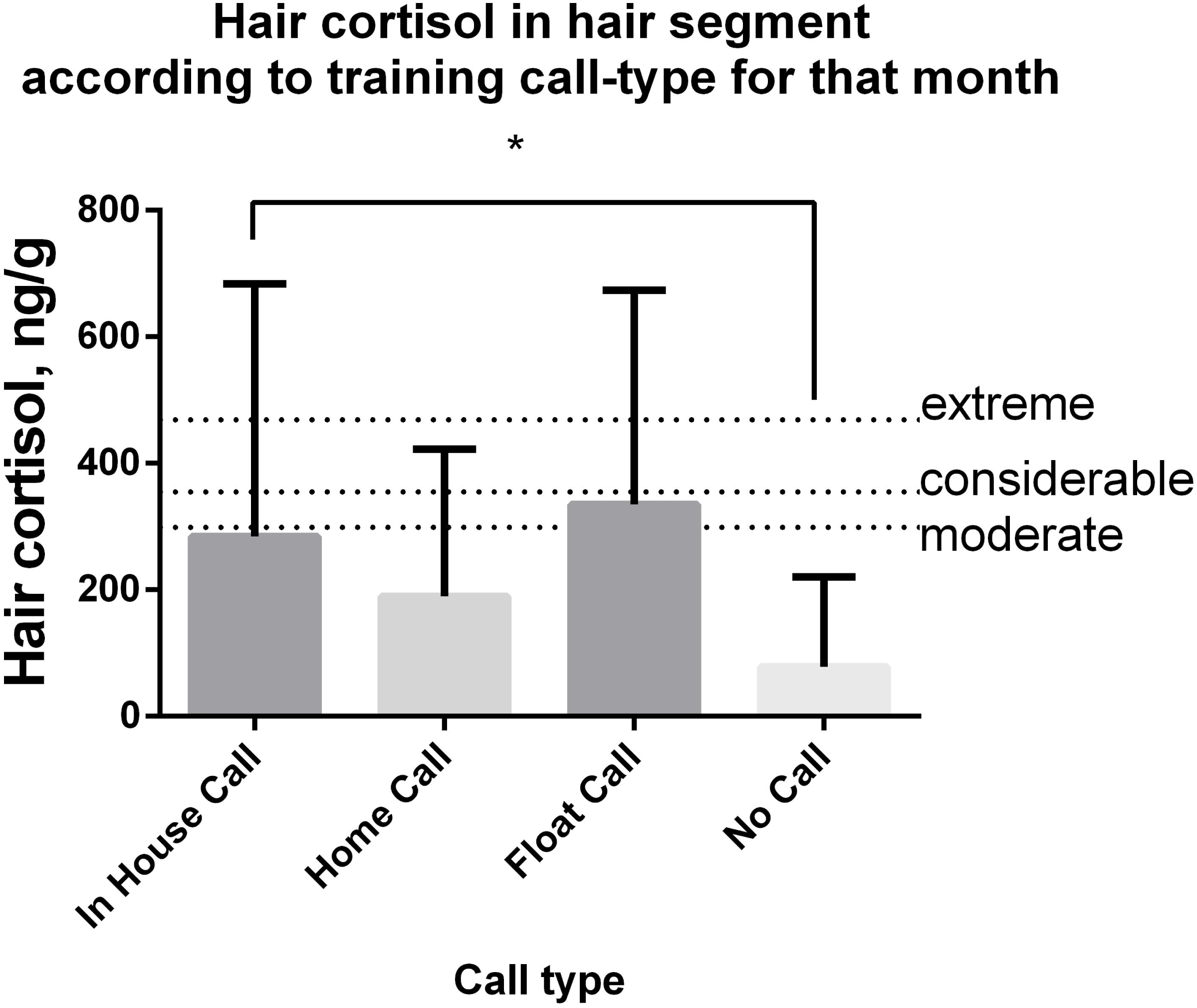
Hair cortisol concentration in relation to call type. Note that moderate elevation is mean +1.5SD (>298.6ng/g), considerable is mean +2SD (>355.4ng/g) and extreme is more than 3 SD above the mean (>468.9 ng/g). * indicates p<0.05 in comparison.

**Table 1:** Sleep interruption (self reported) per block based on call type.

| Call type | Sleep while on call, h, med (IQR) | Est. # pages per night, med (IQR) | Sleep while not on call, h, med (IQR) | Shifts/block med (IQR) | Global call sleep dep index med (IQR) |
| --- | --- | --- | --- | --- | --- |
| In hospital | 2 (1.0-3.0) | 10 (8-15) | 7 (7-8) | 6 (5-7) | 23 (15-29) |
| Home | 5 (3-6) | 5 (4-8) | 8 (7-8) | 5 (4-7) | 6 (0-12) |
| Float | 0.5 (0-2) | 10 (9-15) | 6 (5-7) | 7 (3-7) | 33 (14-42) |
| None |  |  | 6 (5-7) |  |  |

## Discussion

Increased HPA axis activation has been documented in medical trainees. A study by Coeck *et al.* in 1991 of eight internal medicine residents found significantly higher ACTH and cortisol levels at the end of a 24 hour call shift *vs.* a control night when they slept in hospital (1). However, while these measurements reflect acute physiologic stress, HCC offers a long term and retrospective look at chronic HPA axis activation (11). A previous study of medical students (n=55) documented an association with adverse study conditions and HCC, though a control or reference standard for HCC was not provided (29). Mayer *et al.,* in a prospective study of 74 medical interns (equivalent to first year residency) in Germany demonstrated significant HCC increase at two and four months after starting internship, returned to baseline at months six to ten, and rose again from months ten to twelve (30). The authors postulate that these temporal trends reflect working in a novel setting at the beginning of internship and anticipation of the next internship year (30). There was no correlation between self-reported hours of sleep or work hours and HCC, however, no information about call structure was provided.

Our study is different from previous studies measuring temporal changes in HCC in medical trainees (1, 29) through our collection of data about call structure and estimated sleep quantity. Additionally, we report participant level HCC data and utilize published reference data to interpret our results. We found that 40% of trainees experienced at least one month of severe (more than 3SD above the literature-reported mean for a general population) HPA axis activation over a three-month period of training as defined by HCC. In addition, 17% of trainees experienced sustained severe elevation in hair cortisol across three consecutive months of residency. For comparison, using the same laboratory approach to hair cortisol measures, Thomson *et al.* showed that the median hair cortisol level in six patients with Cushing Syndrome was 679 (279-2500) ng/g – similar to the 3-month levels we found in nearly one-fifth of residents (31). A study describing HCC in non-combatant Libyans in war-time (2011) using the same laboratory and assay as our study, reported a median HCC of 186 (63-771) ng/g, lower than that seen among the in-hospital call months for medical residents (27).

Prior studies have showed higher HCC in shift workers compared with day workers (15). Further, a cross-sectional study by Zhang *et al* showed an association between high HCC (defined as third quartile and above) and prevalence of sleep disorders (32). We postulate that sleep deprivation and disruption secondary to in-hospital call contributes to concomitant and sustained HPA axis activation in medical trainees. Though Mayer *et al* did not find HCC to be correlated with sleep, the average self-reported hours of sleep duration in their study (6.6) was notably higher than in ours (0.5 on night float, two on in-hospital call, five on home call and six on no-call) (30), suggesting a lesser degree of sleep disruption and deprivation in their study setting. However, we acknowledge that additional factors such as adjusting to medical training and working in a healthcare environment (33) likely contribute to HPA activation. We therefore cannot draw a definitive causative link between call type and HCC, particularly since in-hospital call and night float are for higher acuity clinical rotations. It is possible elevations in HCC were unrelated to residency stressors, but there were no major life events on the Perceived Stress Scale or Life Stress Inventory that persisted for more than one month.

Our findings have important implications for medical trainees’ long term mental and metabolic health, as HCC has been correlated with burnout symptomatology in health care workers(33), dyslipidemia (34), and elevated BMI (15) in shift workers. In recent decades, appeals for duty hour regulation of medical trainees has led to many residency programs shifting from traditional in-hospital call to a night float system (35, 36, 37). However, it is controversial whether night float improves sleep quality or quantity *vs.* in-hospital call, as studies on this topic have shown conflicting findings (36,38, 39, 40). As we did not demonstrate differences in HCC for night float vs. in-hospital call, our results further call into question whether night float systems are having the intended benefits for the health of trainees. A potential future application of our findings could be the optional, consented monitoring of HCC by trainees by their program directors to complement subjective reported experience. This may help identify trainees at high risk of health consequences of training circumstances and provide an opportunity to intervene through offering psychological support and/or reducing duty hours.

This study has several limitations. We relied on self-report for call type, number of pages received, and hours of sleep, which is subject to recall bias, although self-reporting was done during each rotation segment and not via distant recall. The median age of our participants was 29, and it is well accepted that many stressful life events can happen in this season of life (41), so unmeasured factors unrelated to call or medical training may have been present. We did not have any non-physician or practicing physician controls, so we cannot necessarily say that medical trainees fare worse than staff physicians, other healthcare workers or other shift workers. We do not have baseline (prior to medical training) HCC data, therefore it remains possible that medical training programs are enriched with individuals who have other predispositions to chronic high HPA axis output.

Future studies on this topic should explore whether there are individual factors that protect from or predispose to chronic HPA activation and whether such activation translates to other adverse biomarkers of health such as weight gain, dyslipidemia or hypertension. Additionally, longer-term follow-up analysis of HCC (i.e. beyond residency training) could inform whether these changes persist or dampen over time.

## Conclusion

We have illustrated a high prevalence of chronic hypercortisolism via long-term HPA activation measured through HCC during medical training. For different call models, higher HCC was seen during rotations where residents completed call in the hospital. Furthermore, a high proportion of trainees had severe elevation of HCC during at least one clinical rotation. This study may have important implications for potential long-term health of medical trainees and raises questions about the optimal structure of duty hours and the sequence of care acuity blocks within residency training programs.

## Data Availability

Study data may be made available upon reasonable request and ethical approval

## Acknowledgements

Special thanks to Drs Michael Rieder, Stan VanUum, and Abdelbaset Elzagallaai at Western University for processing of the hair samples and expert input on hair cortisol methodology.

## References

1. Coeck C, Jorens PG, Vandevivere J, Mahler C. ACTH and cortisol levels during residency training. N Engl J Med. 1991;325(10):738.

2. Ferraùa F, Korbonitsb M. Metabolic Syndrome in Cushing’s Syndrome. Front Horm Res. 2018 Apr 5;49:85–103.

3. Nieman LK. Hypertension and cardiovascular mortality in patients with Cushing syndrome. Endocrinology and Metabolism Clinics. 2019 Dec 1;48(4):717–25.

4. Webb SM, Valassi E. Quality of life impairment after a diagnosis of Cushing’s syndrome. Pituitary. 2022 Oct;25(5):768–71.

5. Findling JW, Raff H. Recognition of nonneoplastic hypercortisolism in the evaluation of patients with cushing syndrome. Journal of the Endocrine Society. 2023 Aug;7(8):bvad087.

6. Vaccarino V, Bremner JD. Stress and cardiovascular disease: an update. Nature Reviews Cardiology. 2024 Sep;21(9):603–16.

7. Charmandari E, Tsigos C, Chrousos G. Endocrinology of the stress response. Annu. Rev. Physiol.. 2005 Mar 17;67:259–84.

8. Jutla SK, Yuyun MF, Quinn PA, Ng LL. Plasma cortisol and prognosis of patients with acute myocardial infarction. Journal of Cardiovascular Medicine. 2014 Jan 1;15(1):33–41.

9. Tang AR, Rabi DM, Lavoie KL, Bacon SL, Pilote L, Kline GA. Prolonged hypothalamic-pituitary-adrenal axis activation after acute coronary syndrome in the GENESIS-PRAXY cohort. European Journal of Preventive Cardiology. 2018 Jan 1;25(1):65–72.

10. Morris MC, Rao U, Garber J. Cortisol responses to psychosocial stress predict depression trajectories: social-evaluative threat and prior depressive episodes as moderators. Journal of affective disorders. 2012 Dec 20;143(1-3):223–30.

11. Wester VL, van Rossum EF. Clinical applications of cortisol measurements in hair. Eur J Endocrinol. 2015;173(4):M1–10.

12. Russell E, Koren G, Rieder M, Van Uum S. Hair cortisol as a biological marker of chronic stress: current status, future directions and unanswered questions. Psychoneuroendocrinology. 2012;37(5):589–601.

13. Stalder T, Tietze A, Steudte S, Alexander N, Dettenborn L, Kirschbaum C. Elevated hair cortisol levels in chronically stressed dementia caregivers. Psychoneuroendocrinology. 2014;47:26–30.

14. Wosu AC, Valdimarsdóttir U, Shields AE, Williams DR, Williams MA. Correlates of cortisol in human hair: implications for epidemiologic studies on health effects of chronic stress. Ann Epidemiol. 2013;23(12):797–811.e2.

15. Manenschijn L, van Kruysbergen RG, de Jong FH, Koper JW, van Rossum EF. Shift work at young age is associated with elevated long-term cortisol levels and body mass index. J Clin Endocrinol Metab. 2011;96(11):E1862–5.

16. Basner M, Asch DA, Shea JA, Bellini LM, Carlin M, Ecker AJ, et al. Sleep and Alertness in a Duty-Hour Flexibility Trial in Internal Medicine. N Engl J Med. 2019;380(10):915–23.

17. Garcia-Leon, M. A., Peralta-Ramirez, M. I., Arco-Garcia, L., Romero-Gonzalez, B., Caparros-Gonzalez, R. A., Saez-Sanz, N., Santos-Ruiz, A. M., Montero-Lopez, E., Gonzalez, A., & Gonzalez-Perez, R. (2019). Hair cortisol concentrations in a Spanish sample of healthy adults. PLoS One, 14(8), e0221883.

18. Bolster L, Rourke L. The Effect of Restricting Residents’ Duty Hours on Patient Safety, Resident Well-Being, and Resident Education: An Updated Systematic Review. J Grad Med Educ. 2015;7(3):349–63.

19. Moeller A, Webber J, Epstein I. Resident duty hour modification affects perceptions in medical education, general wellness, and ability to provide patient care. BMC Med Educ. 2016;16:175.

20. Ahmed N, Devitt KS, Keshet I, Spicer J, Imrie K, Feldman L, et al. A systematic review of the effects of resident duty hour restrictions in surgery: impact on resident wellness, training, and patient outcomes. Ann Surg. 2014;259(6):1041–53.

21. Ahmed N, Devitt KS, Keshet I, Spicer J, Imrie K, Feldman L, et al. A systematic review of the effects of resident duty hour restrictions in surgery: impact on resident wellness, training, and patient outcomes. Ann Surg. 2014;259(6):1041–53.

22. Lockley SW, Cronin JW, Evans EE, Cade BE, Lee CJ, Landrigan CP, et al. Effect of reducing interns’ weekly work hours on sleep and attentional failures. N Engl J Med. 2004;351(18):1829–37.

23. Veasey S, Rosen R, Barzansky B, Rosen I, Owens J. Sleep loss and fatigue in residency training: a reappraisal. JAMA. 2002;288(9):1116–24.

24. Cohen S, Kamarck T, Mermelstein R. A global measure of perceived stress. Journal of health and social behavior. 1983 Dec 1:385–96.

25. Holmes TH, Rahe RH. The social readjustment rating scale. Journal of psychosomatic research. 1967.

26. Greff, MJ, et al. Hair cortisol analysis: An update on methodological considerations and clinical applications. Clinical Biochemistry. 2019; 63: 1–9.

27. Etwel F, Russell E, Rieder MJ, Van Uum SH, Koren G. Hair cortisol as a biomarker of stress in the 2011 Libyan War. Clin Invest Med 2014;37(6):e403–8.

28. Igboanugo S, O’Connor C, Zitoun OA, Ramezan R, Mielke JG. A systematic review of hair cortisol in healthy adults measured using immunoassays: Methodological considerations and proposed reference values for research. Psychophysiology. 2024;61(1):e14474.

29. Heming M, Angerer P, Apolinario-Hagen J, Nater UM, Skoluda N, Weber J. The association between study conditions and hair cortisol in medical students in Germany - a cross-sectional study. J Occup Med Toxicol. 2023;18(1):7.

30. Mayer SE, Lopez-Duran NL, Sen S, Abelson JL. Chronic stress, hair cortisol and depression: A prospective and longitudinal study of medical internship. Psychoneuroendocrinology. 2018;92:57–65.

31. Thomson S, Koren G, Fraser LA, Rieder M, Friedman TC, Van Uum SH. Hair analysis provides a historical record of cortisol levels in Cushing’s syndrome. Experimental and Clinical Endocrinology & Diabetes. 2010 Feb;118(02):133–8.

32. Zhang Y, Shen J, Zhou Z, Sang L, Zhuang X, Chu M, et al. Relationships among shift work, hair cortisol concentration and sleep disorders: a cross-sectional study in China. BMJ Open. 2020;10(11):e038786.

33. Marcil MJ, Cyr S, Marin MF, Rosa C, Tardif JC, Guay S, et al. Hair cortisol change at COVID-19 pandemic onset predicts burnout among health personnel. Psychoneuroendocrinology. 2022;138:105645.

34. Zhu L, Zhang Y, Song L, Zhou Z, Wang J, Wang Y, et al. The relationships of shift work, hair cortisol concentration and dyslipidaemia: a cohort study in China. BMC Public Health. 2022;22(1):1634.

35. Fletcher KE, Underwood W, 3rd, Davis SQ, Mangrulkar RS, McMahon LF, Jr., Saint S. Effects of work hour reduction on residents’ lives: a systematic review. JAMA. 2005;294(9):1088–100.

36. Chowdhary A, Davis JA, Ding L, Taravati P, Feng S. Resident Sleep During Traditional Home Call Compared to Night Float. J Acad Ophthalmol (2017). 2023;15(2):e204–e8.

37. Aggarwal S, Wisely CE, Gross A, Challa P. Transition to a Night Float System in Ophthalmology Residency: Perceptions of Resident Wellness and Performance. J Acad Ophthalmol (2017). 2022;14(1):e120–e6.

38. Kelly-Schuette K, Shaker T, Carroll J, Davis AT, Wright GP, Chung M. A Prospective Observational Study Comparing Effects of Call Schedules on Surgical Resident Sleep and Physical Activity Using the Fitbit. J Grad Med Educ. 2021;13(1):113–8.

39. Ko JS, Readal N, Ball MW, Han M, Pierorazio PM. Call Schedule and Sleep Patterns of Urology Residents Following the 2011 ACGME Reforms. Urol Pract. 2016;3(2):147–52.

40. Cavallo A, Ris MD, Succop P. The night float paradigm to decrease sleep deprivation: good solution or a new problem? Ergonomics. 2003;46(7):653–63.

41. Demiray B, Gulgoz S, Bluck S. Examining the life story account of the reminiscence bump: why we remember more from young adulthood. Memory. 2009;17(7):708–23.

